# A simplified model for the analysis of COVID-19 evolution during the lockdown period in Italy

**DOI:** 10.1101/2020.06.02.20119883

**Authors:** Roberto Simeone

## Abstract

A simplified model applied to COVID-19 cases detected and officially published by the italian government [1], seems to fit quite well the time evolution of the disease in Italy during the period feb-24th - may-19th 2020.

The hypothesis behind the model is based on the fact that in the lockdown period the infection cannot be transmitted due to social isolation and, more generally, due to the strong protection measures in place during the observation period. In this case a compartment model is used and the interactions between the different compartments are simplified. The sample of cases detected is intended as a set of individuals susceptible to infection which, after being exposed and undergoing the infection, were isolated (’treated’) in such a way they can no longer spread the infection.

The values obtained are to be considered indicative.

The same model has been applied both to the data relating to Italy and to some regions of Italy (Lombardia, Piemonte, Lazio, Campania, Calabria, Sicilia, Sardegna), generally finding a good response and indicatively interesting values (see chap. 5).

The only tuning parameter is the ‘incubation period’ *τ* that, together with the calculated growth rate *κ* of the exponential curve used to approximate the early stage data.

**Conclusions:** A simplified compartmental model that uses only the incubation period and the exponential growth rate as parameters is applied to the COVID-19 data for Italy in the lockdown period finding a good fitting.

**Revision History:** This section summarize the history of revisions.

*Revision # 1:*

- Errata corrige in section 1 (Introduction): the equations that summarize the relationship between the parameters were wrong. This revised version contains the correct equations at page 2.
- The synchronization criteria is updated. No need to use a threshold different to the one used to determine the growth coefficient. The results are now updated with the synchronization point near to the 20% of the maximum value of the cases detected per day: 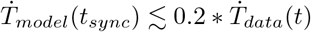
- Modifications in section 4 (Model results for Italy). It is appropriate to use an exponential function instead of a logistic function to find the growth rate in the initial phase. Section 4 and the results are now updated.
- Some non-substantial corrections in the descriptive part.

*Revision # 2:*

- Errata corrige in the system differential equation 6: in the the derivative of S were reported a wrong additional term N. Now the equation 6 is correct.

*Revision # 3:*

- New approach to detect the exponential rate and new concept for the transfer coefficients.
- **Exponential rate:** The old criteria was oriented to the growth of the cases: *y*_Δ_*_t_* = *y*_0_ * *e^k^*^Δ^*^t^* thus: *y*_0_ + Δ*y* = *y*_0_ * *e^k^*^Δ^*^t^*. The exponential growth rate was then: *k* = *log*(1 + Δ*y*/*y*_0_)/Δ*t*. The new criteria is oriented to the growth of the differences Δ*y* = *e^k^*^Δ^*^t^* − 1 obtaining: *k* = *log*(1 + Δ*y*)/Δ*t*.
- **Transfer coefficients:** The new approach is based on the following assumptions: *α_SE_* = *ke^kδ^* [*day*^−1^]: this coefficent is supposed to be the variation of the exponential growth per unit of time (*δ* = 1 *day*). *α_EI_* = 1/*τ* [*day*^−1^] where τ is the incubation period (this assumption is not changed). *α*_IT_ = *kδ/τ* [*day*^−1^] this coefficent is supposed to be proportional to the ratio *kδ*/*τ*. The constant *δ* =1 [*day*] represent the unit variation in time.
- **Basic reproduction number:** With the above assumptions, the basic reproduction number become: 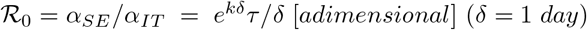
- **Revision summary:** The old approach, although adapting well to the data, presented several inconsistencies in the parameters, and in particular on the relationship between 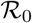 and *k*. In this revision the new approach still shows a good fit to the data and shows congruent relationships between the parameters.

## 1 Introduction

After the so-called ‘phase 1’, characterized by social distancing and a general halt of all ‘non-essential’ activities, the curve of the new COVID-19 cases detected daily dropped sharply. This period, also called ‘lockdown’ started with the decree of the Italian government of March 9th and lasted until May 4th 2020.

The data of the daily detections of infected cases are available to everyone, starting from February 24th, 2020, on the website [1]. This allowed a comparison of the growth curve of the cases detected with a compartmental model of the Susceptible-Exposed-Infected-Recovered type (see [2], [4], [3],) which provides the temporal evolution of the number of individuals belonging to the different compartments. The model has been suitably simplified to take into account the lockdown situation.

The period under consideration (from Feb 24th to May 19th 2020) is sufficiently limited to allow us to exclude the effects of births and deaths on the population, therefore in this context we consider a constant population of *N* individuals, with *N* = *S* + *E* + *I* + *R*.

In the initial phase, the spread of the infection was not controlled, thus allowing an exponential increase in the number of Exposed and Infected. Between March 4th and 9th drastic measures were taken which, starting from Lombardia, were extended to all italian regions. The measures in the blocking period aim to limit the spread of the infection as much as possible by isolating people in homes and, where it is not possible (essential services), imposing individual protections and disinfection of common areas.

The lockdown period, simplifying, can also be thought of as a type of treatment for infective individuals. We can therefore imagine that infective individuals, detected daily, in general are managed (’treated’) in such a way that they cannot infect other susceptible individuals. This ‘treatment’ includes both hospitalization and isolation.

The lockdown period, characterized by strong social isolation, also allows us to neglect any potential transfers between the various departments beyond the basic path *S* → *E* → *I* → *T*.

To avoid confusing the SEIR model with the lockdown approximation, in this context, we will call T (Treated) the compartment of infected people who are no longer able to transmit the pathogen.

This ‘SEIT’ simplified model is based on the assumption that the transfer coefficient from S to T is the variation of the exponential growth in the unit of time, and the transfer coefficient from I to T is directly proportional to the growth rate and inversely proportional to the incubation period. Finally the transfer coefficient from E to I is the reciprocal of the incubation period.

The model seems to fit well with the data relating to the Italian regions.

## 2 SEIR compartmental models

The mathematical models for the study of the spread of infections are based on a subdivision of the population into ‘compartments’ and on systems of differential equations to represent their temporal evolution. The most common models are the SEIR models (Susceptible-Exposed-Infective-Recovered) described in [2], [4], [3].

In the SIR and SEIR models the variables are represented by the number of individuals in the different compartments over time (in this case on a daily basis).

*S* (*Susceptible*) Number of uninfected individuals susceptible to pathogen infection.

*E* (*Exposed*) Number of individuals who have been exposed to the pathogen but who, throughout the incubation period, are not spreading the infection.

*I* (*Infected*) Number of individuals infected and capable of transmitting the pathogen to others.

*R* (*Recovered*) Number of individuals who have passed the infection and are no longer able to transmit the pathogen to others.

The evolution over time of the variables S, E, I, R can be modeled (see eq.1) considering the interactions between the different compartments and taking into due consideration the relationships that contribute to the increase the number of infected than those that only describe the displacements between the various compartments without increasing the *I* variable.

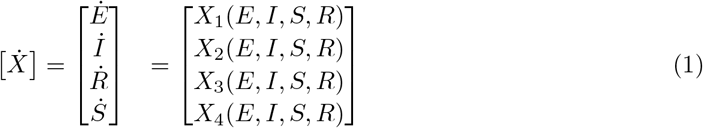

In the system of differential equations (1) the components *X_i_* are functions of the variables (*E*, *I*, *R*, *S*) and 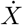 their derivative over time.

The solution of (1) represents the evolution over time of the number of individuals in the different sectors.

Given a situation of equilibrium (DFE: Disease Free Equilibrium) represented by a set of values *X*_0_, the stability of the system can be studied by analyzing the contribution of infectious components close to the DFE state.

By construction, the *S* and *R* compartments do not generate infections, therefore the variables to be considered in the stability study near DFE are *Ė* and *İ* (2).

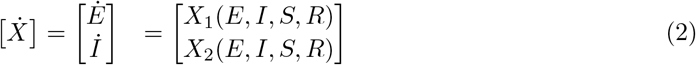

In order to study the stability near a DFE, it’is important to distinguish the ‘new infections’ from the movement between the departments, the differential equation (2) associated with each variable is divided into two components:

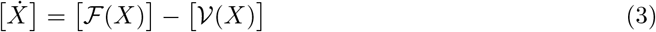

By construction 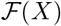 and 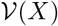 represent:

- 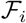: new infections.
- 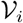: transfers between compartments only.

From this representation, given the initial conditions *S*_0_, *E*_0_, *I*_0_, *R*_0_, and the interaction parameters between the various compartments that characterize 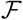 and 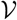, solving the (1) the temporal evolution of the number of individuals in the different compartments is obtained.

To study the stability of the system near equilibrium conditions (ref. [2]) we consider a DFE as solution of the differential equations (1) and the perturbative impact of the two infectious variables using the two Jacobian matrices of size 2 * 2 relating to *I* and *E* (see eq. 2).

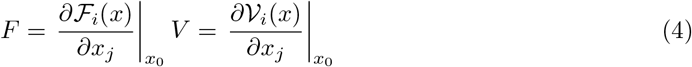

The product *FV*^−1^ is known as ‘next generation matrix’ [2]. Each element 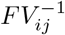 represents the number of ‘secondary’ infections of the *i* compartment due to a single infected of the *j* compartment assuming that the whole population is susceptible. Each element of the matrix is a ‘reproduction number’ which indicates how much an infectious person in one compartment affects the number of infected in another compartment.

The dominant eigenvalue (spectral radius) of the ‘next generation matrix’ *FV*^−1^ is known as the ‘basic reproduction number’ 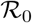 and represents the number of new cases generated, on average, from a single case during its infectious period in a completely susceptible set of individuals. 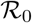 also represents the stability threshold of the system near an equilibrium state. A value of 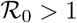 implies that the the system will move away from a state of equilibrium, with consequent spreading of the infection (see [2], [4])

- 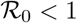: the equilibrium (DFE) is locally stable and diffusion does not extend;
- 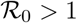: the equilibrium is unstable allowing the spread of the infection.

## 3 A simplification (SEIT) to model the lockdown

As already mentioned, we will consider lockdown as a form of treatment of the infection since all members of the *I* sector, also as a result of the lockdown measures and, more generally, of social isolation, distancing and individual protection, reduce their ability to spread the pathogen. In this case, to avoid confusion we will call *T* (Treated) the *R* (Recovered) variable of the SEIR model.

We also will simplify the interactions between compartments by considering only the forward ones *S* → *E* → *I* → *T* and we will consider the total number of individuals *N* = *S* + *E* + *I* + *T* constant. In this context, *N* represents a sample of individuals, with homogeneous distribution, susceptible to infection in an environment with constant transfer coefficients between compartments.

The transfers between compartments in this case are:

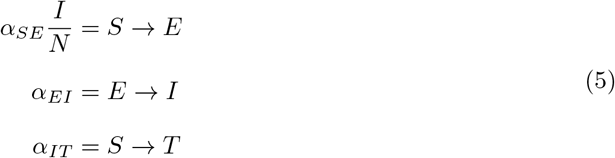

We note that:

- The transfer *S* → *E* is proportional to the product of the ratio of infected individuals (*I*/*N*) by the daily contact coefficient *α_SE_* of the individuals in *S*.
- The coefficient *α_EI_* represents the daily passage of individuals from E to I. The value 1/*α_EI_* is basically related to the incubation time of the infection.
- the value *α_IT_* basically represents the speed of ‘treatment’ of the individuals in the compartment *I*, so that they can no longer transmit the pathogen, with consequent transfer to compartment *T*.

Recalling that, in this context, we consider *N* constant over time, we have that *Ṡ*+*Ė*+*İ*+*Ṫ* = 0, and the system of differential equations results:

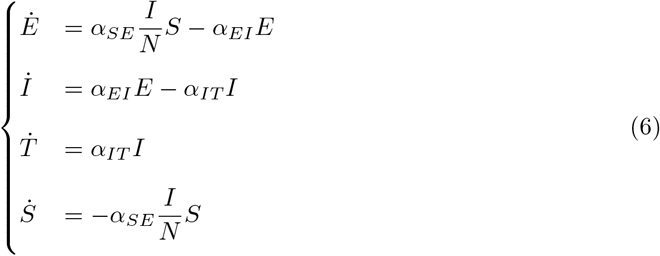

Where the total number of individuals is *N* = *S* + *E* + *I* + *T* and the variables are ordered so that the first two are the ‘infectious variables’ (*E*, *I*).

To study the stability we will consider *Ė* and *İ* and wh have:

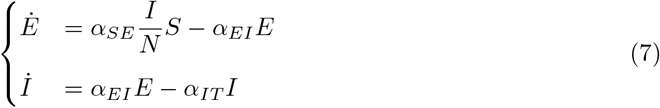

in eq.(7) *α_SE_*(*I*/*N*)*S* it is the only element that contributes to the increase of *I*.

Considering the separation of variables in two groups ‘infective’ and ‘transfer’ (see eq. 3), the eq. (7) in vector terms can be written as:

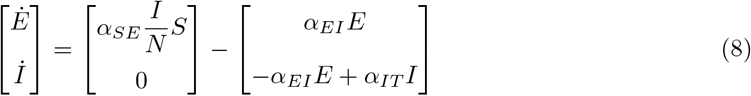

A trivial solution of equilibrium DFE is had for *S*_0_ = *N* which corresponds to the initial vector:

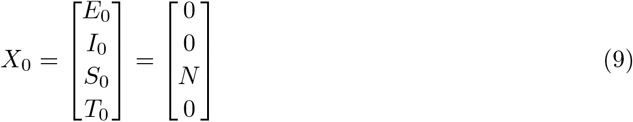

The Jacobian matrices (see eq. 4) near the equilibrium condition *X*_0_ (*S* = *N*) for the infectious variables *E* and *I* are therefore:

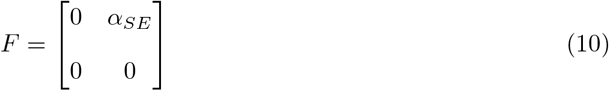

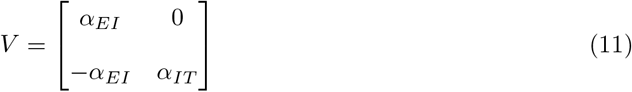

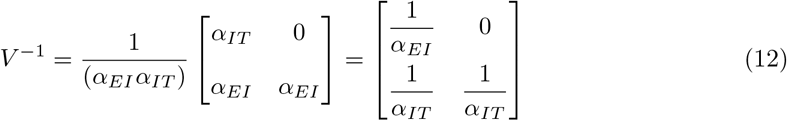

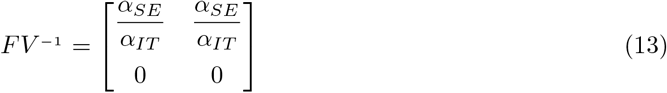

The eigenvalues of the next generation matrix *FV*^−1^ are:

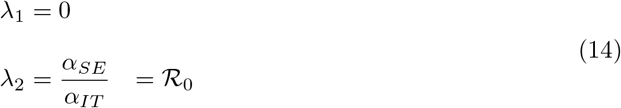

The greater eigenvalue (spectral radius) *λ*_2_ represents the ‘basic reproduction number’ 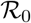.

In this simplified representation it is easy to see that, once the equilibrium condition (DFE) is disrupted, 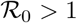 implies that the daily contacts rate *α_SE_* in S compartment, that will produces new infected, is greater than the number of infective individuals treated per day *α_IT_*. In this case, on average, an infected individual produce more than one new infected during his infectious period. As consequence the system moves away from equilibrium by extending the spread of the pathogen.

On the other hand, if *α_SE_* < *α_IT_* 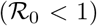 then the number of new infected per day is less than the neutralization rate and the infection will not spread further.

## 4 Model results for Italy

The DFE equilibrium represented by the trivial solution (9) was disrupted (15) with an individual ‘Exposed’ within a sample of *N* individuals. All *S*_0_ = *N* − 1 individuals are susceptibles to infection.

*N* is equal to the total number of cases detected in Italy from 24 Feb to May 19th [1]: *N* = 226699 individuals.

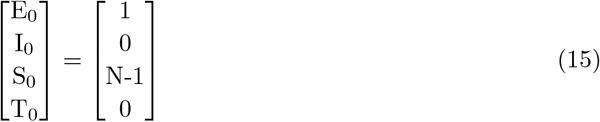

In this context we consider the data [1] as detected values (*T_data_*) for the compartment *T*. In order to smooth the effect of the variability of the number of cases between the various regions, an initial disruption based on the percentage of cases detected should be carried out. As already mentioned the results are indicative, then, for simplicity, the perturbation described by (15) is used for all simulations.

In order do estimate the parameters for this model, an exponential growth rate must be detected from the available data.

Because we are looking for a growth in terms of differences we can assume:

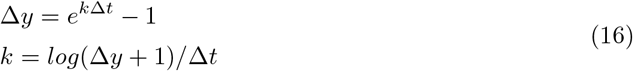

where Δ*y* is the difference of cases detected in the period Δ*t*.

The first problem to be addressed, for the definition of the model, is the estimation of the parameters *α_SE_*, *α_EI_*, *α_IT_* for the transfer rate of individuals between the compartments. The following assumptions have been made:

- in the early phase the exponential growth can be approximated using an exponential curve. To estimate the exponential rate *k* we used for Δ*y* (16) the difference from the total cases detected at the day corresponding to the maximum daily difference during the early growth and the cases detected at the first day (see fig. 1a);
- the *α_SE_* rate is is assumed to be *α_SE_* = *ke^kδ^* corresponding to the derivative of Δ*y* (16) and using *δ* =1 day.
- the parameter *α_EI_* is the reciprocal of the incubation time *τ* which will be the only tuning parameter of the model: *α_EI_* = 1/*τ*.
- the parameter *α_IT_* is assumed to be directly proportional to the exponential growth per unit of time and inversely proportional to the incubation period *τ*, then *α_IT_* = *kδ/τ*.

**Figure 1:**
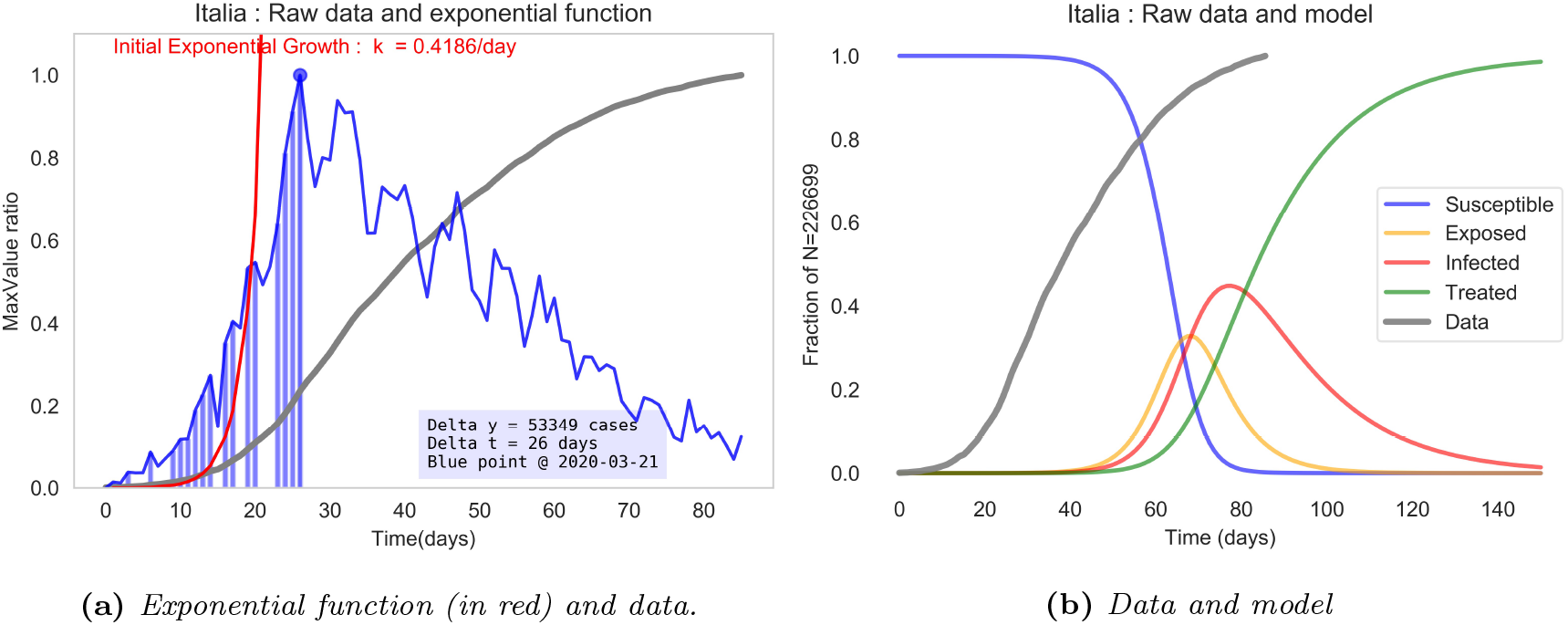
(a) Exponential function (red) used to approximate the data in the initial phase and daily variations of T_data_ (blue). (b) Response of the model to the perturbation eq. (15) near the DFE. The ordinate scales indicate in (a) the ratio with respect to the maximum values and in (b) the ratio on the total cases N of the number of individuals in the compartments E, I, S, T. In gray (T_data_) are plotted the values of the COVID-19 cases detected in Italy until may 19th 2020: [1]).

With the above assumptions, and recalling that *δ* =1 *day*, the following relationships have been identified:

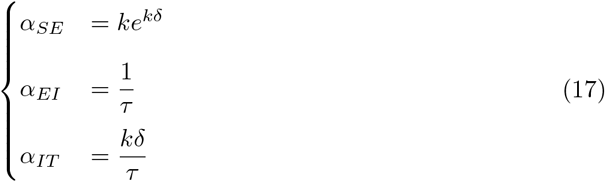

In this context, for 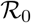 we can write (ref. 14 and 17):

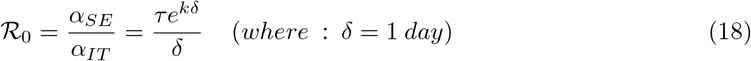

From the available data it was easy to derive *k* (ref. Eq. 16). The only ‘tuning’ parameter is *τ* which has been modified on the basis of a qualitative approximation of the model to the data. Based on these parameters and relationships (17), the simplified SEIT model described above has been set up. The time evolution of the Treated compartment *(T_model_)* of the model has been then compared with the values *T_data_* available for COVID-19 Italia (ref. [1]) finding a good representation with a value of the tuning parameter *τ* = *6days* (see tab. 1 and fig. 1).

The model and the data were ‘synchronized’ on their normalized values at the value used to calculate the growth rate k, obtaining an indicative estimate of the day *t*_0_ in which the diffusion began and of the two peak values for *E* and for *I* (see fig. 2).

**Figure 2:**
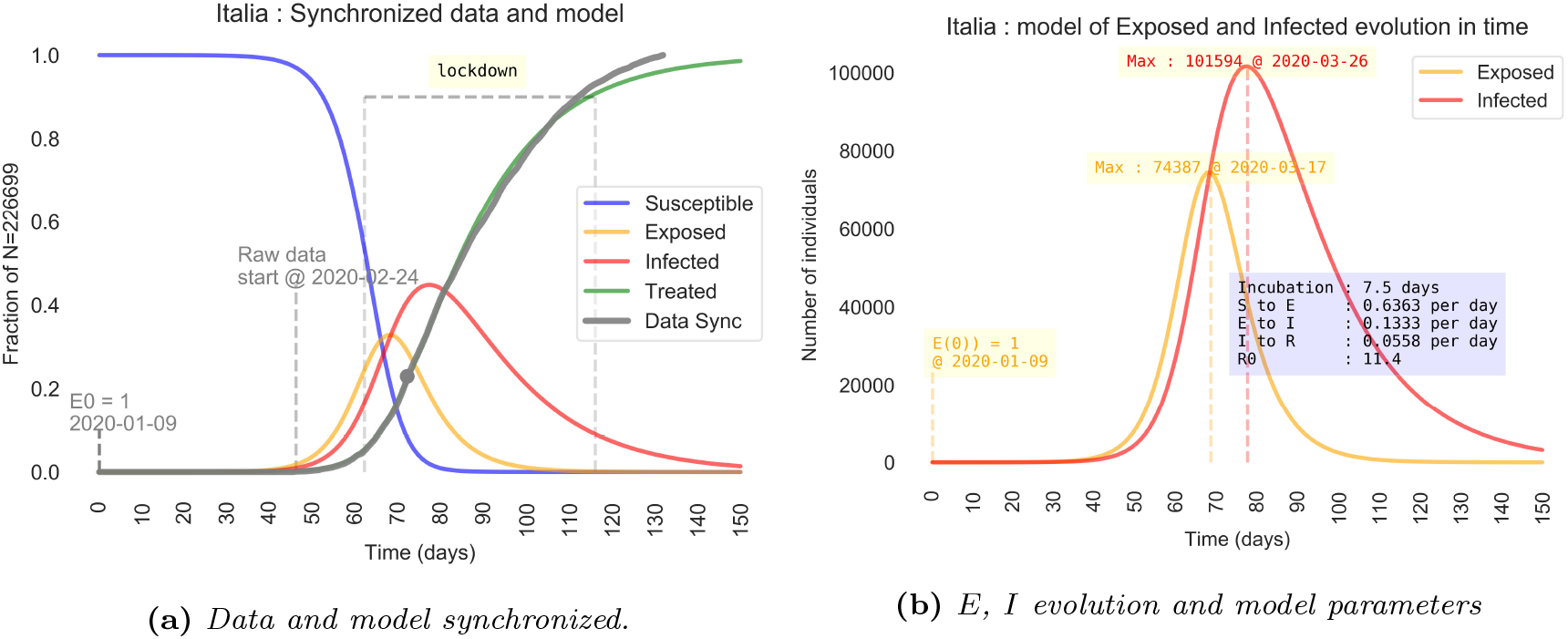
(a) The curves of model (T) and data (T_data_) have been synchronized to the point where k is calculated (the grey point). The ordinate scale is the ratio on the total number of cases N. (b) Detail of time evolution of E and I curves. The ordinate scale is the the number of individuals in the compartments E,I.

The graphs in fig. 3 show the trend of the variation over time of the cases detected and of the compartment *T* (fig. 3a) and the difference between the model and the cases detected (fig. 3b).

**Figure 3:**
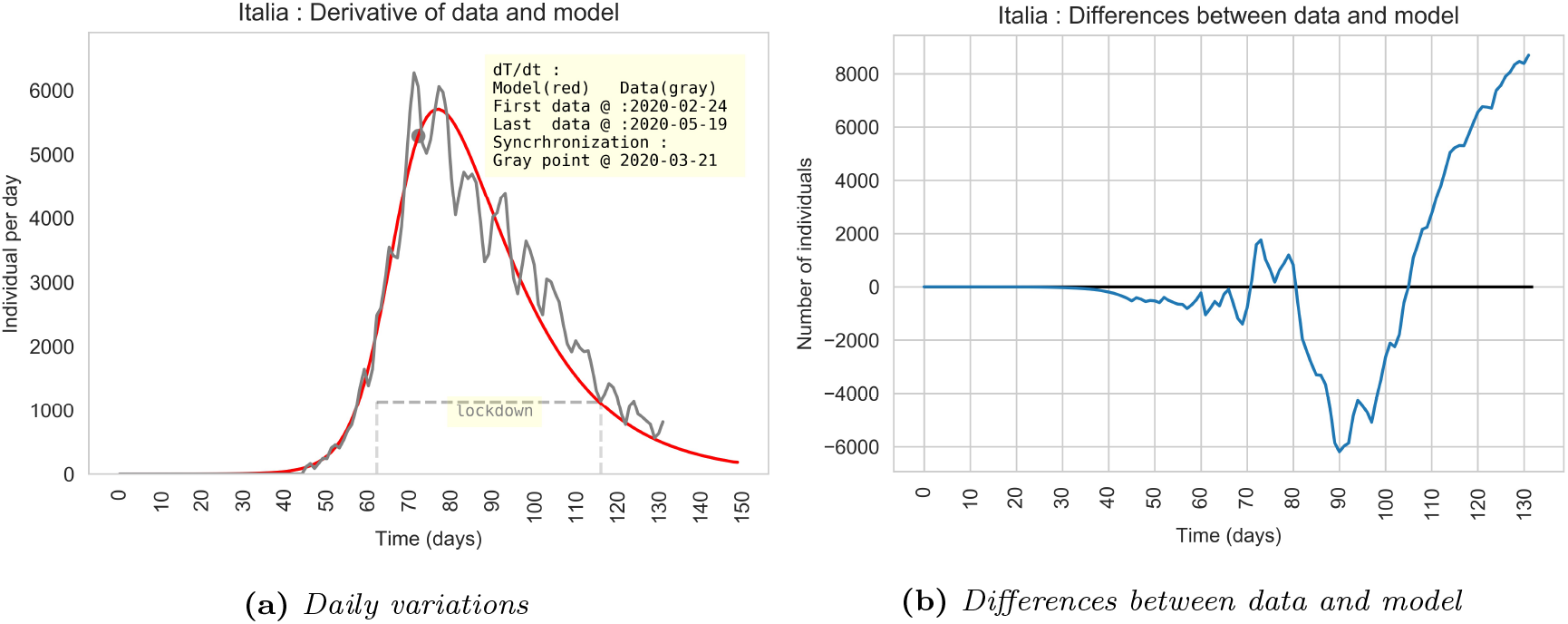
(a) Daily variation of T_data_ and of the T model (red). (b) Differences between T_data_ and the model T.

**Table 1:**
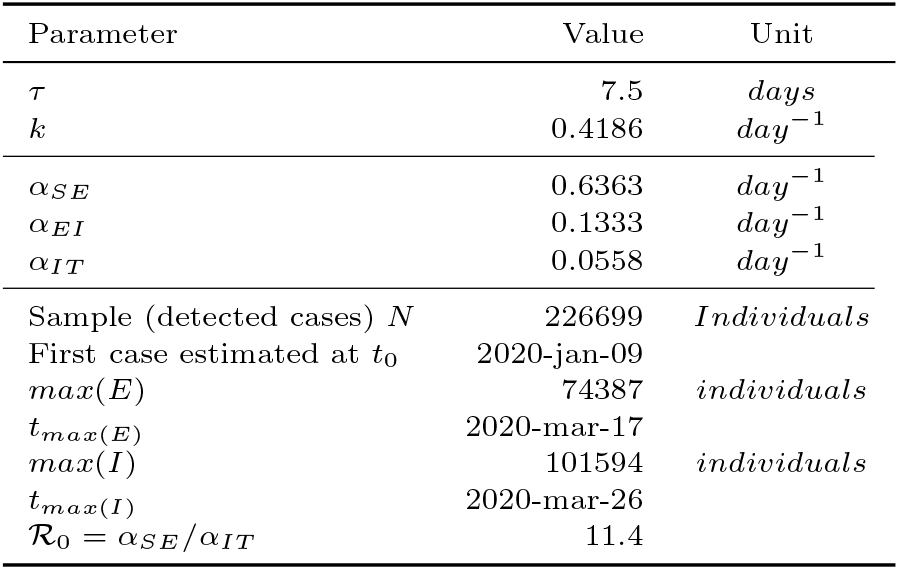
Parameters used and results for Italy. The average incubation period that best fit the data is: τ = 7.5 days. The daily detected data cover a period from Feb 24th to May 19th 2020.

| Parameter | Value | Unit |
| --- | --- | --- |
| $\tau$ | 7.5 | days |
| $k$ | 0.4186 | day<sup>-1</sup> |
| $\alpha_{SE}$ | 0.6363 | day<sup>-1</sup> |
| $\alpha_{EI}$ | 0.1333 | day<sup>-1</sup> |
| $\alpha_{IT}$ | 0.0558 | day<sup>-1</sup> |
| Sample (detected cases) $N$ | 226699 | Individuals |
| First case estimated at $t_0$ | 2020-jan-09 | |
| $max(E)$ | 74387 | individuals |
| $t_{max}(E)$ | 2020-mar-17 | |
| $max(I)$ | 101594 | individuals |
| $t_{max}(I)$ | 2020-mar-26 | |
| $\mathcal{R}_0 = \alpha_{SE}/\alpha_{IT}$ | 11.4 | |

## 5 The simplified model applied to some regions of Italy

Table 2 summarize the application of the simplified model to some italian regions, the graphic details are in the dedicated sections.

**Table 2:** Parameters and results of the model applied, to some regions of Italy. The results are qualitative and must not be understood as valid in absolute terms. The N values are the cumulative COVID-19 cases detected from Feb 24th to May 19th in [1]. In this table t_0_ represents the date of the first case estimated by the model synchronized with the data detected at the value used to estimate the exponential growth rate.

| Region | $\tau$ | $\kappa$ | $\alpha_{SE}$ | $\alpha_{EI}$ | $\alpha_{IT}$ | $N$ | $t_0$ | $Max(E)$ | $t_E$ | $Max(I)$ | $t_I$ | $\mathcal{R}_0$ |
| --- | --- | --- | --- | --- | --- | --- | --- | --- | --- | --- | --- | --- |
| Italia | 7.5 | 0.4186 | 0.6363 | 0.1333 | 0.0558 | 226699 | 2020-01-09 | 74387 | 2020-03-17 | 101594 | 2020-03-26 | 11.4 |
| Lombardia | 8.0 | 0.3900 | 0.576 | 0.1250 | 0.0488 | 85481 | 2020-01-05 | 27852 | 2020-03-13 | 39556 | 2020-03-23 | 11.8 |
| Piemonte | 6.5 | 0.3109 | 0.4242 | 0.1538 | 0.0478 | 29727 | 2020-01-13 | 7365 | 2020-03-23 | 13647 | 2020-04-02 | 8.9 |
| Lazio | 4.0 | 0.2765 | 0.3647 | 0.2500 | 0.0691 | 7505 | 2020-01-23 | 1146 | 2020-03-23 | 2820 | 2020-04-01 | 5.3 |
| Campania | 4.0 | 0.3084 | 0.4199 | 0.2500 | 0.0771 | 4707 | 2020-01-30 | 789 | 2020-03-22 | 1753 | 2020-03-30 | 5.4 |
| Calabria | 3.0 | 0.2565 | 0.3315 | 0.3333 | 0.0855 | 1153 | 2020-02-03 | 120 | 2020-03-24 | 353 | 2020-03-31 | 3.9 |
| Sicilia | 4.0 | 0.2353 | 0.2977 | 0.2500 | 0.0588 | 3403 | 2020-01-16 | 452 | 2020-03-19 | 1294 | 2020-03-29 | 5.1 |
| Sardegna | 4.0 | 0.3330 | 0.4646 | 0.2500 | 0.0832 | 1354 | 2020-02-08 | 242 | 2020-03-21 | 500 | 2020-03-28 | 5.6 |

### 5.1 Lombardia

**Figure 4:**
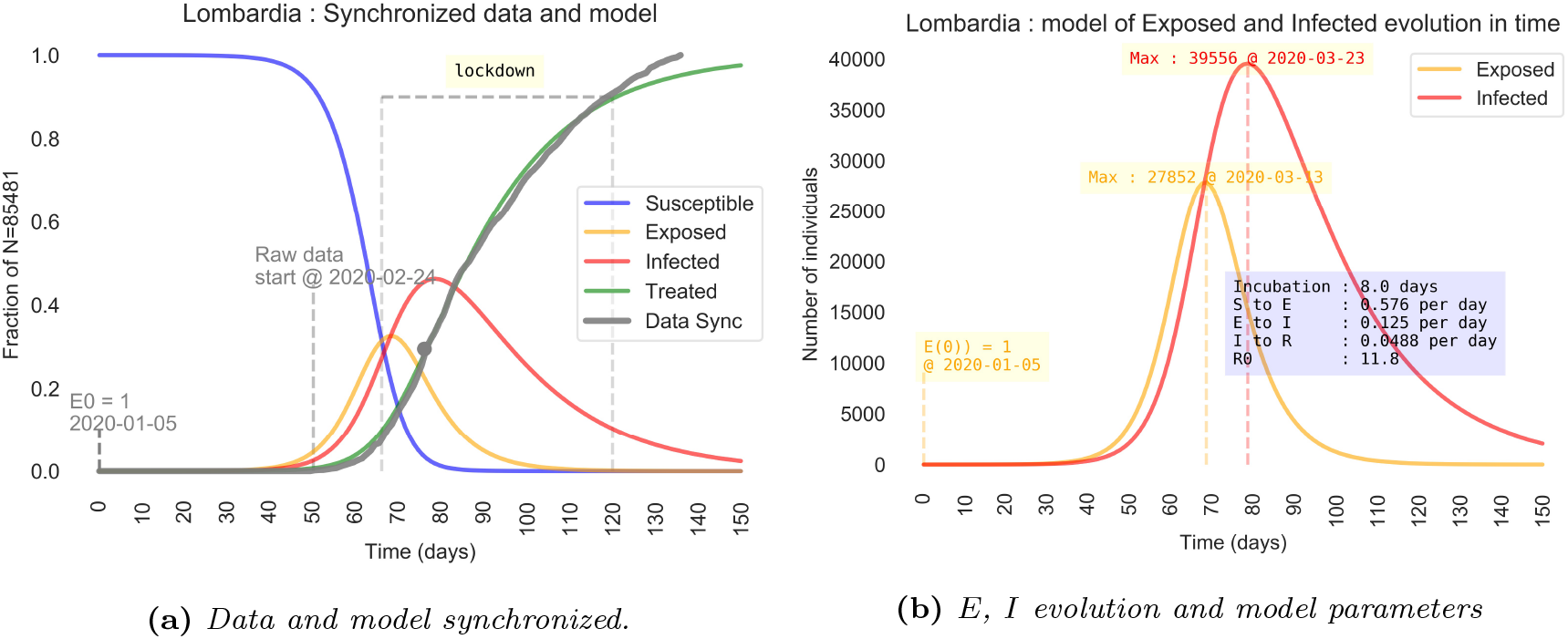
(a) The curve T of the model and T_data_ data has been synchronized at the point where the growth rate has been estimated. The ordinate scale is the ratio on the total number of cases N. (b) Detail of time evolution of E and I curves. The ordinate scale is the the number of individuals in the compartments E,I.

**Figure 5:**
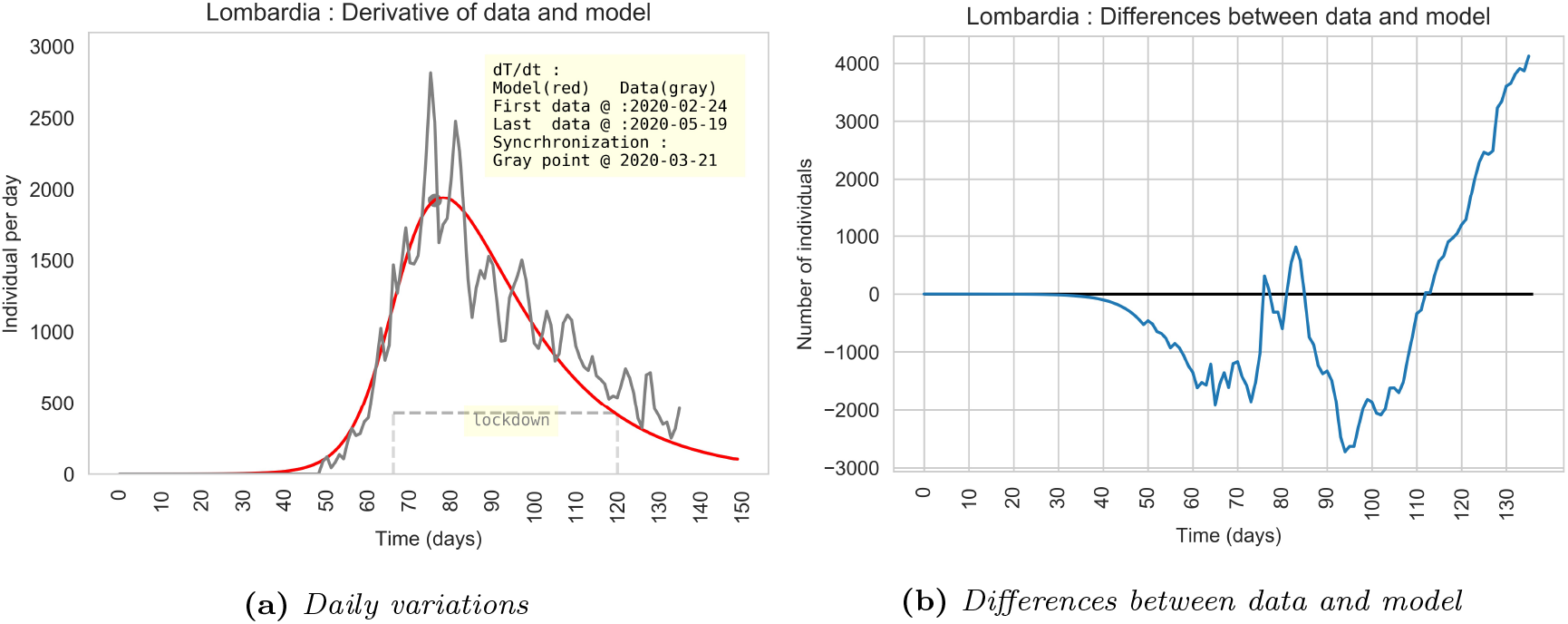
(a) Daily variation of T_data_ and of the T model (red). (b) Differences between T_data_ and the model T.

### 5.2 Piemonte

**Figure 6:**
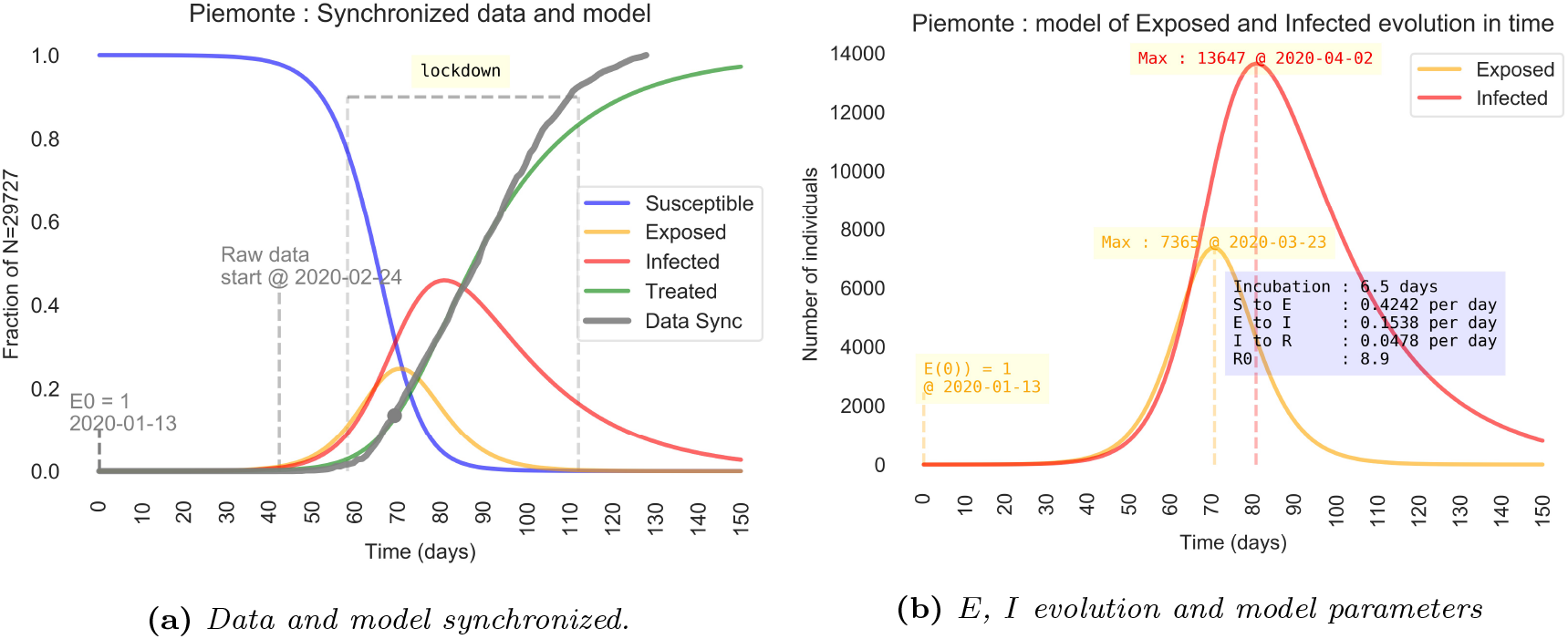
(a) The curve T of the model and T_data_ data has been synchronized at the point where the growth rate has been estimated. The ordinate scale is the ratio on the total number of cases N. (b) Detail of time evolution of E and I curves. The ordinate scale is the the number of individuals in the compartments E,I.

**Figure 7:**
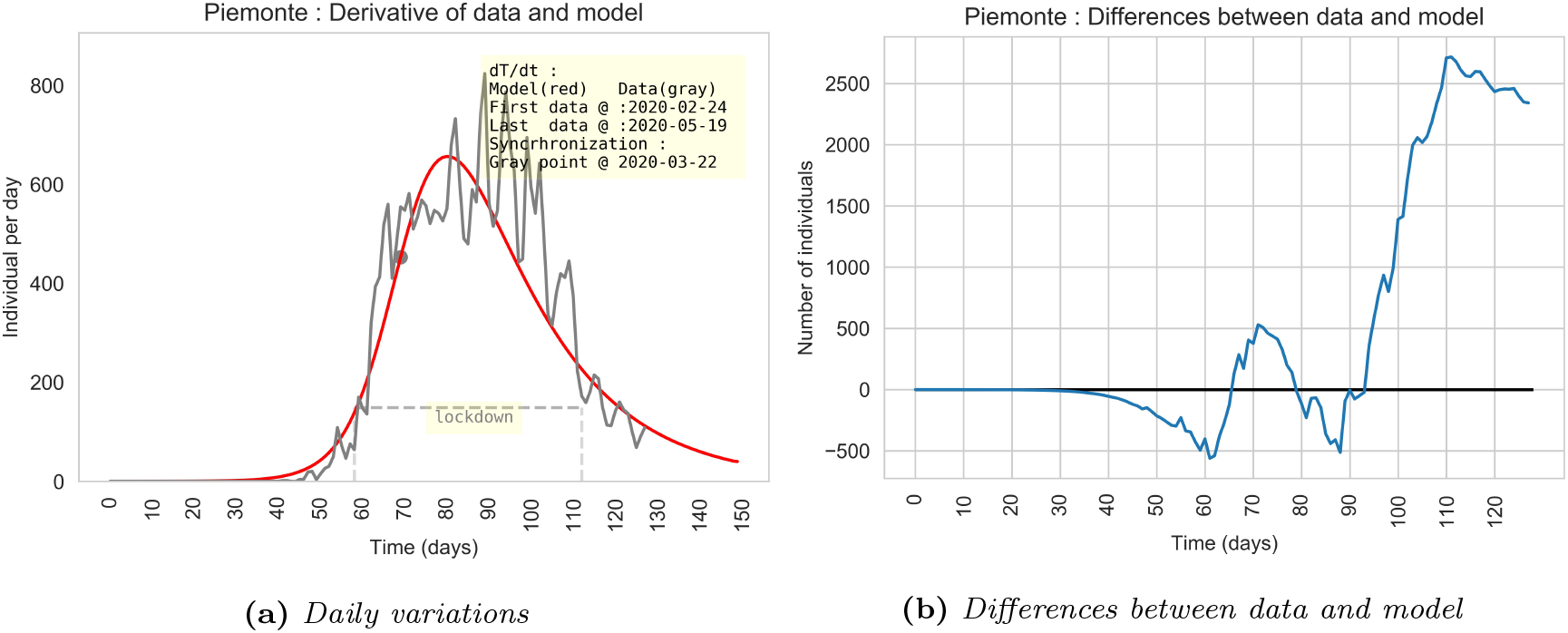
(a) Daily variation of T_data_ and of the T model (red). (b) Differences between T_data_ and the model T.

### 5.3 Lazio

**Figure 8:**
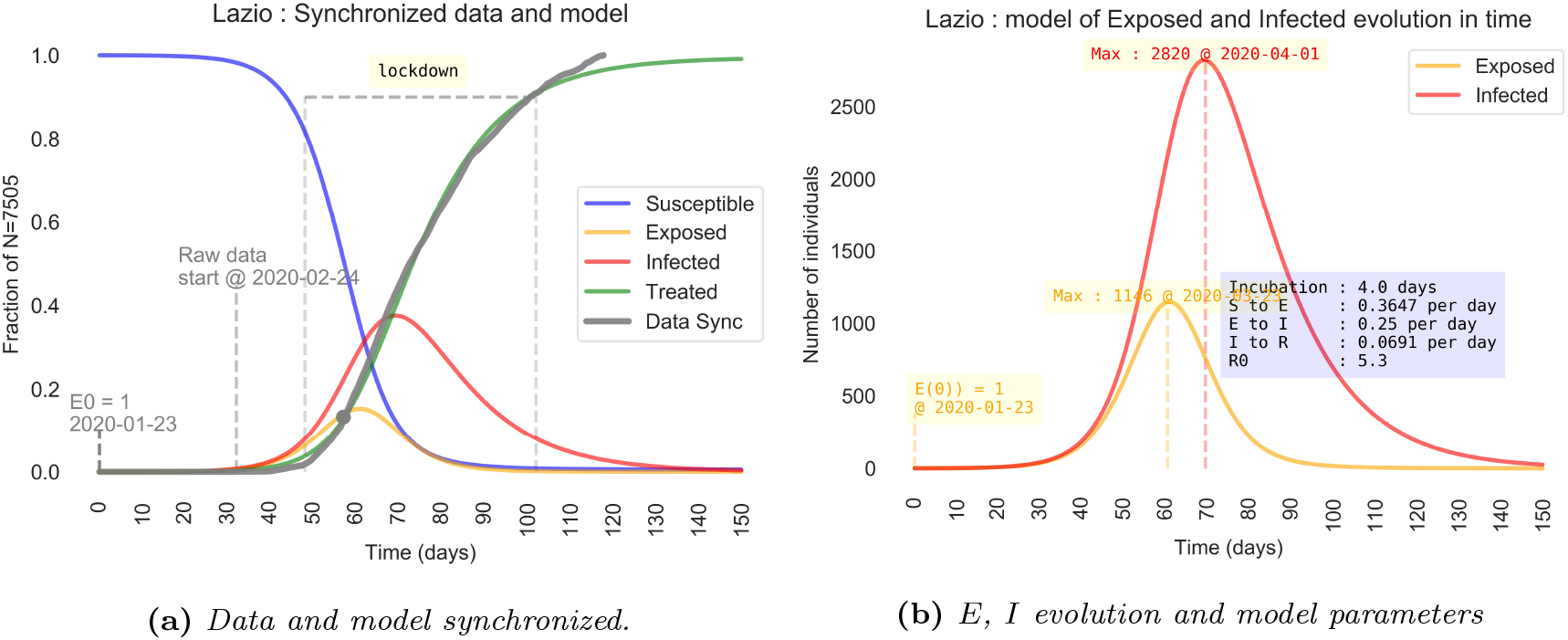
(a) The curve T of the model and T_data_ data has been synchronized at the point where the growth rate has been estimated.The ordinate scale is the ratio on the total number of cases N. (b) Detail of time evolution of E and I curves. The ordinate scale is the the number of individuals in the compartments E,I.

**Figure 9:**
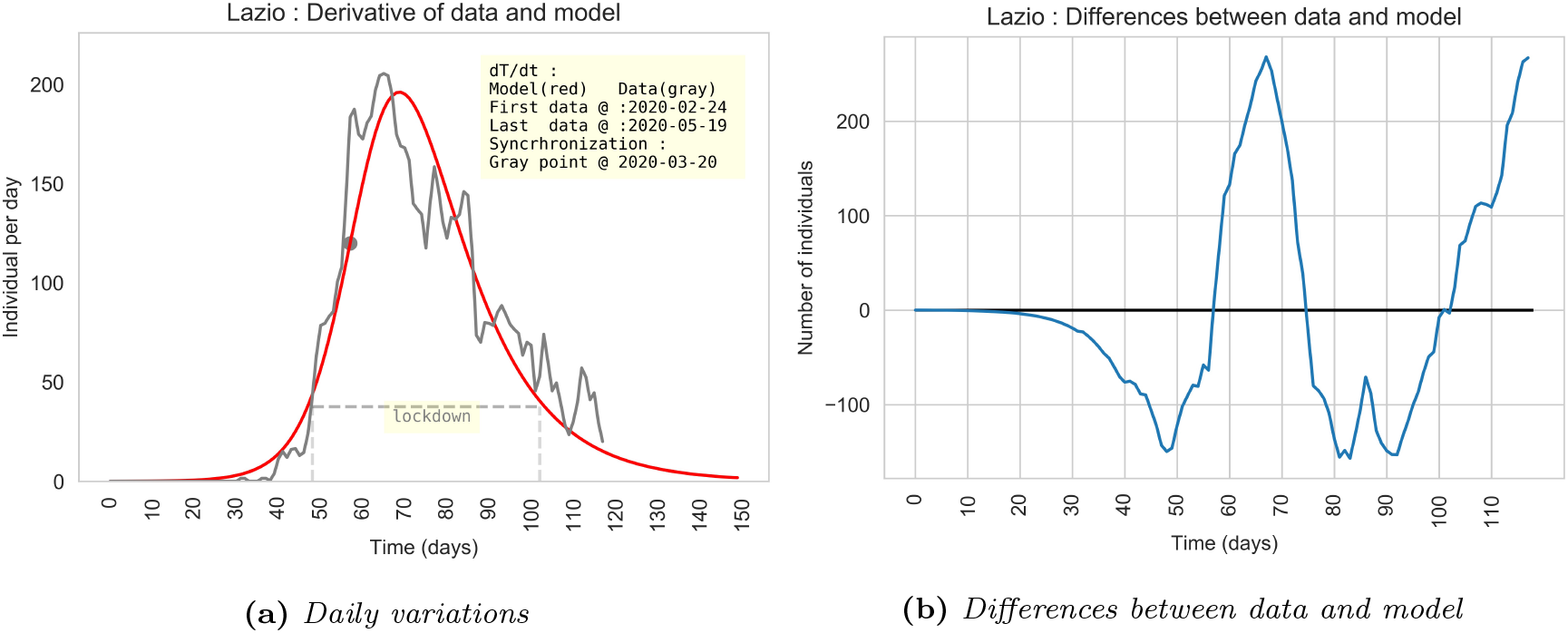
(a) Daily variation of T_data_ and of the T model (red). (b) Differences between T_data_ and the model T.

### 5.4 Campania

**Figure 10:**
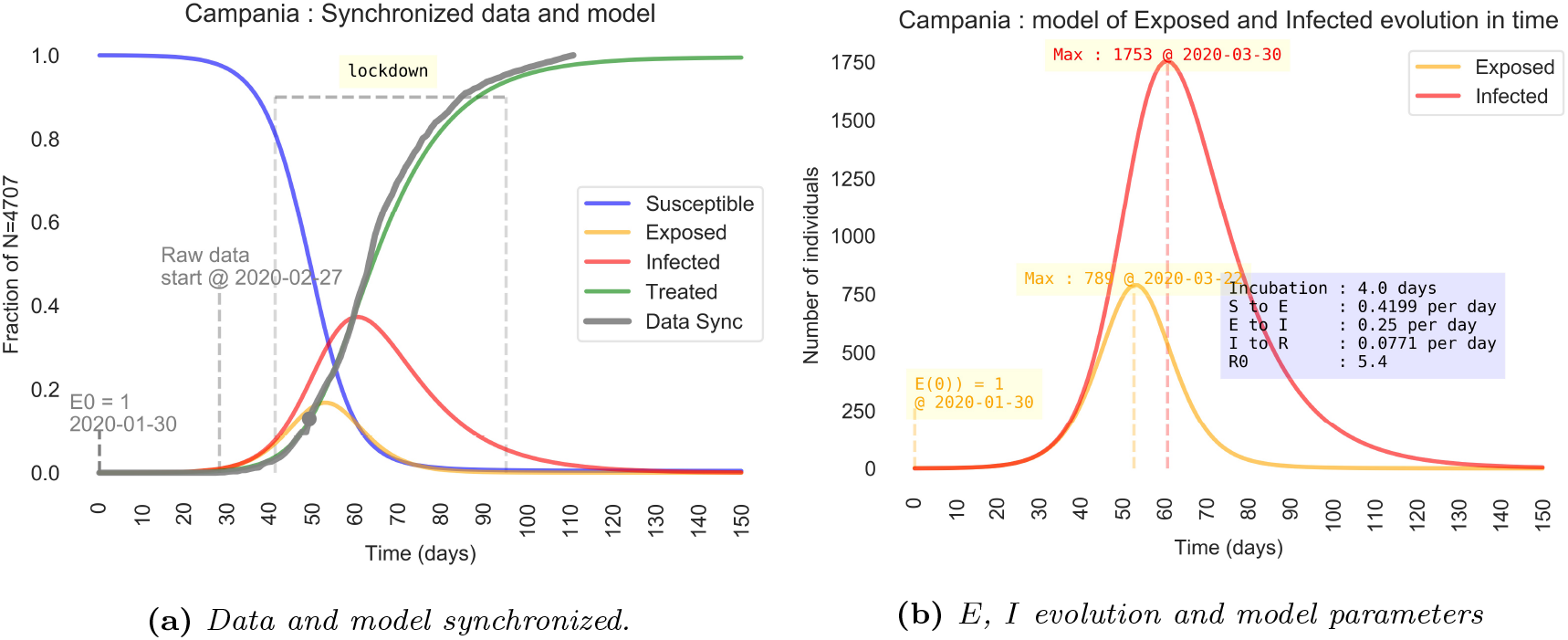
(a) The curve T of the model and T_data_ data has been synchronized at the point where the growth rate has been estimated. The ordinate scale is the ratio on the total number of cases N. (b) Detail of time evolution of E and I curves. The ordinate scale is the the number of individuals in the compartments E, I.

**Figure 11:**
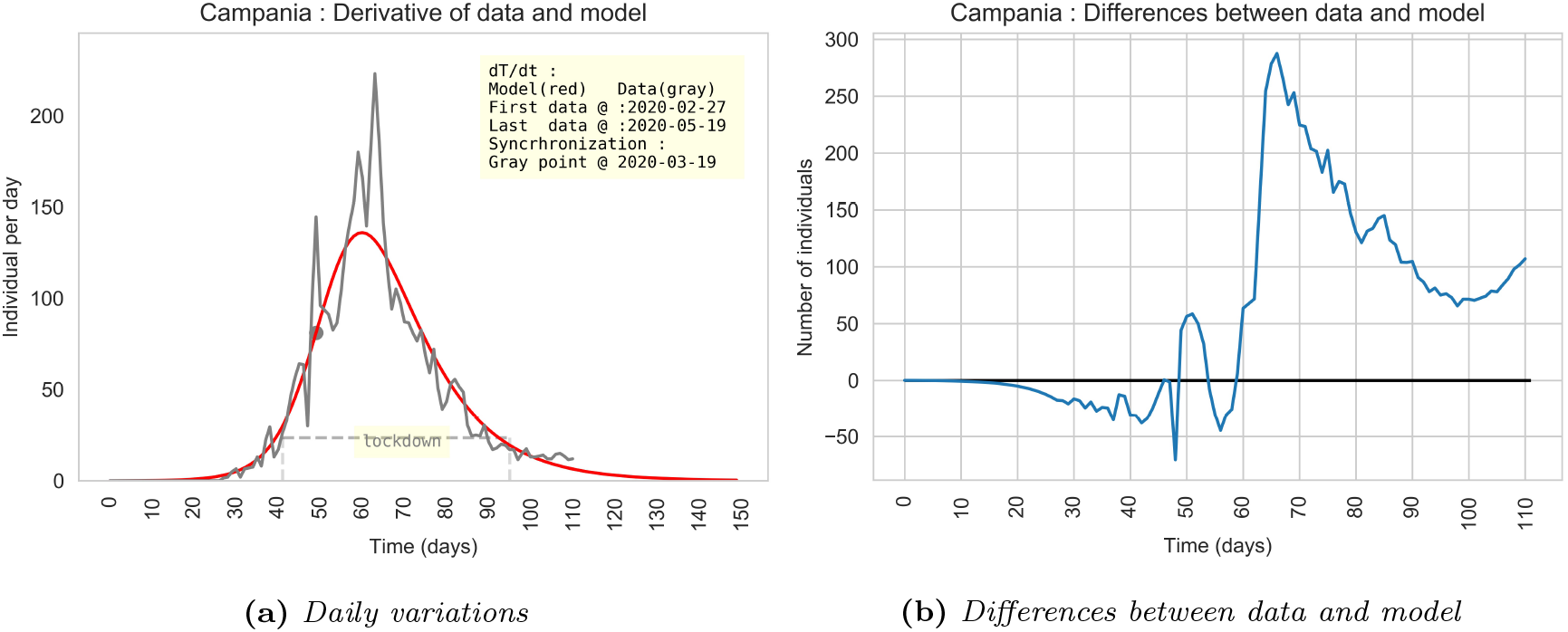
(a) Daily variation of T_data_ and of the T model (red). (b) Differences between T_data_ and the model T.

### 5.5 Calabria

**Figure 12:**
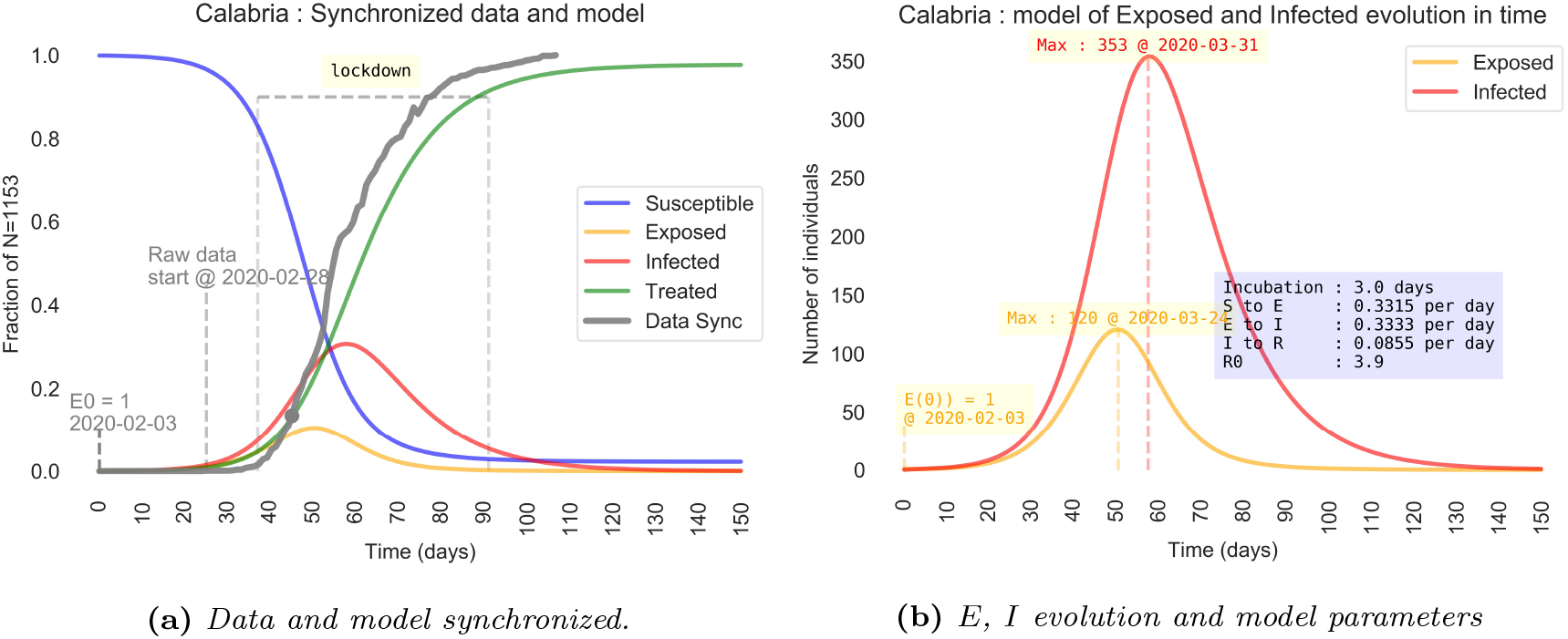
(a) The curve T of the model and T_data_ data has been synchronized at the point where the growth rate has been estimated. The ordinate scale is the ratio on the total number of cases N. (b) Detail of time evolution of E and I curves. The ordinate scale is the the number of individuals in the compartments E, I.

**Figure 13:**
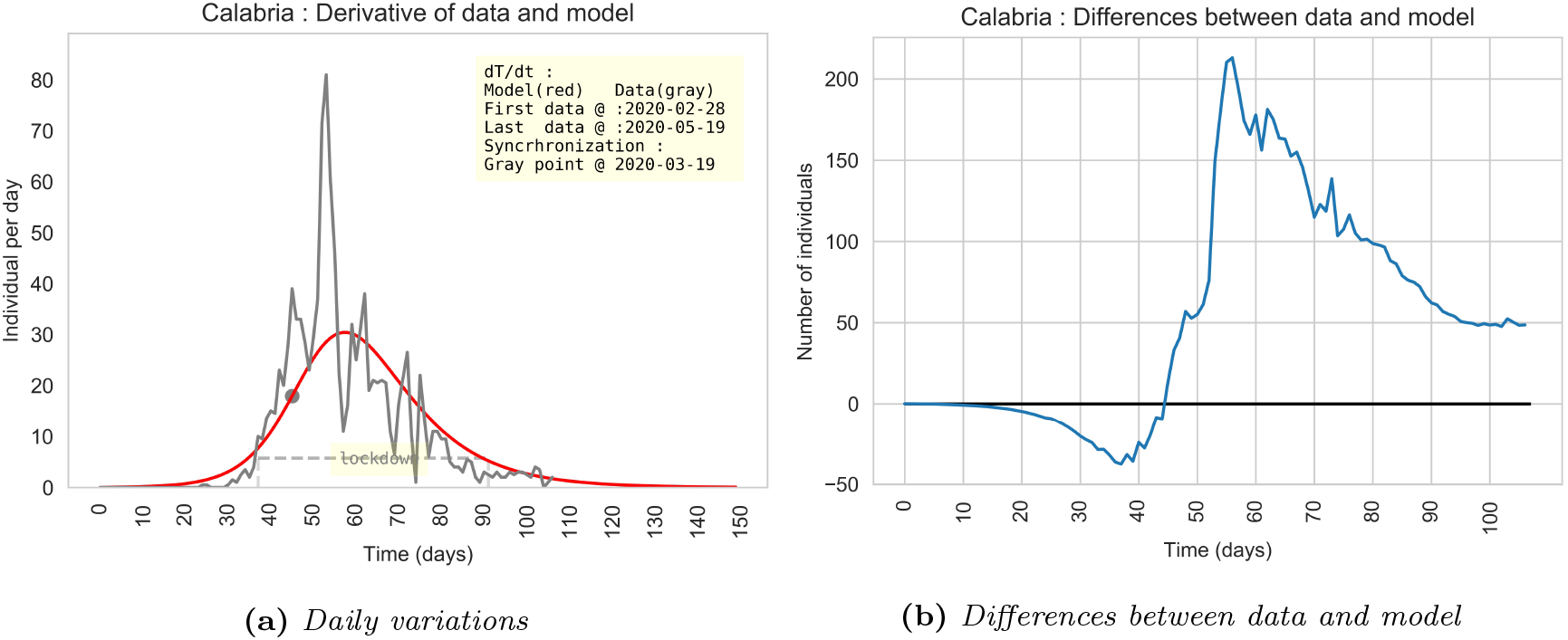
(a) Daily variation of T_data_ and of the T model (red). (b) Differences between T_data_ and the model T.

### 5.6 Sicilia

**Figure 14:**
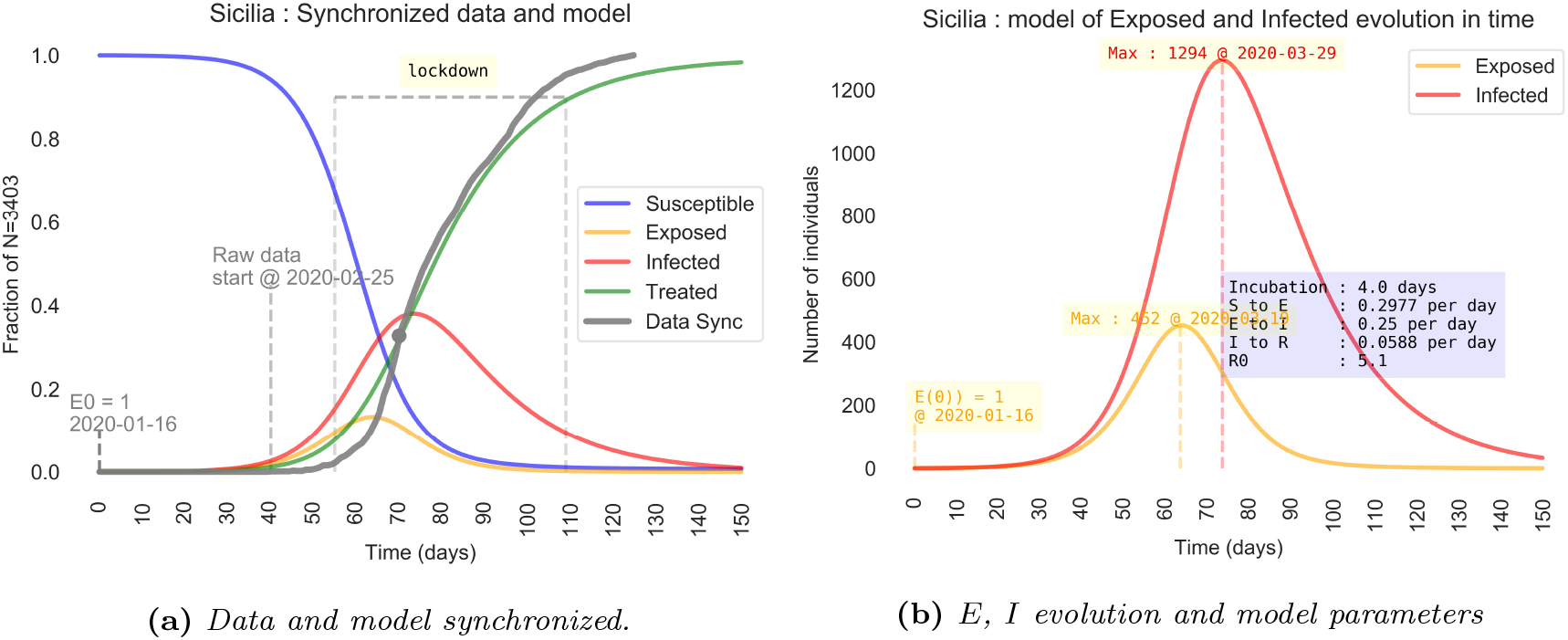
(a) The curve T of the model and T_data_ data has been synchronized at the point where the growth rate has been estimated. The ordinate scale is the ratio on the total number of cases N. (b) Detail of time evolution of E and I curves. The ordinate scale is the the number of individuals in the compartments E, I.

**Figure 15:**
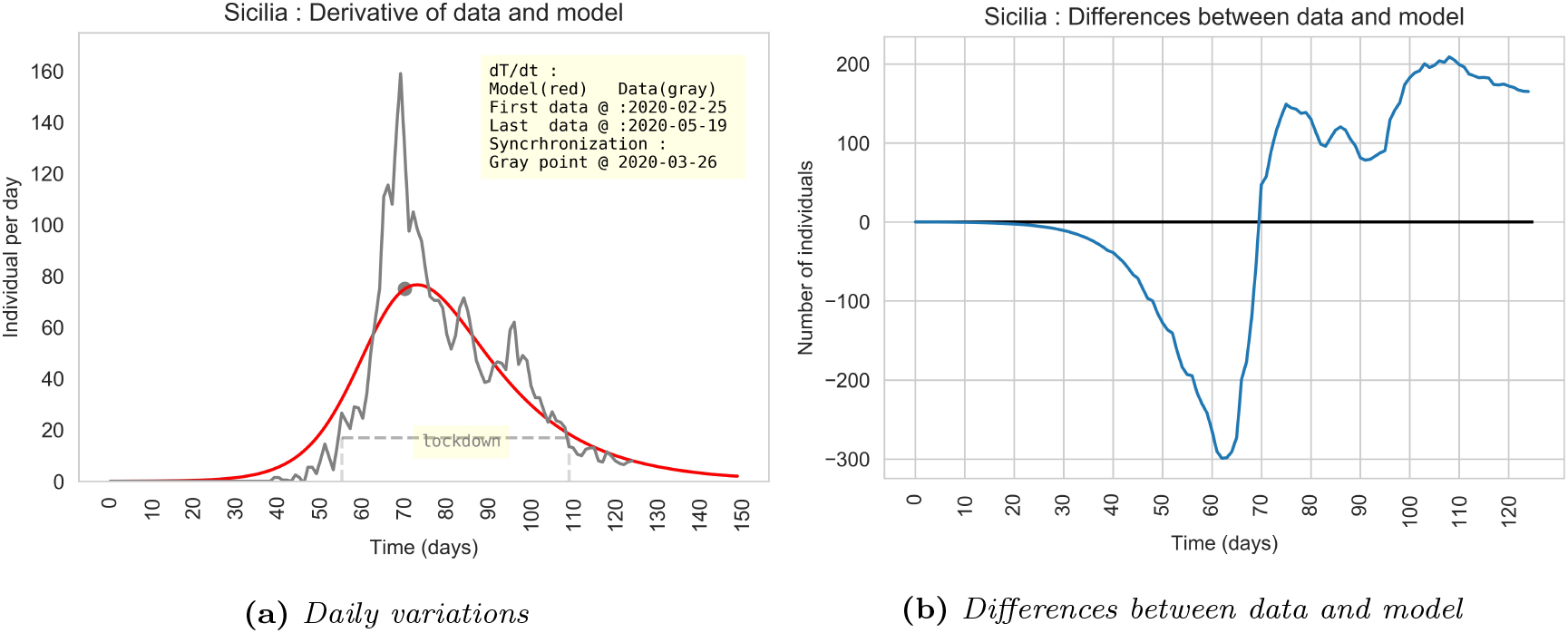
(a) Daily variation of T_data_ and of the T model (red). (b) Differences between T_data_ and the model T.

### 5.7 Sardegna

**Figure 16:**
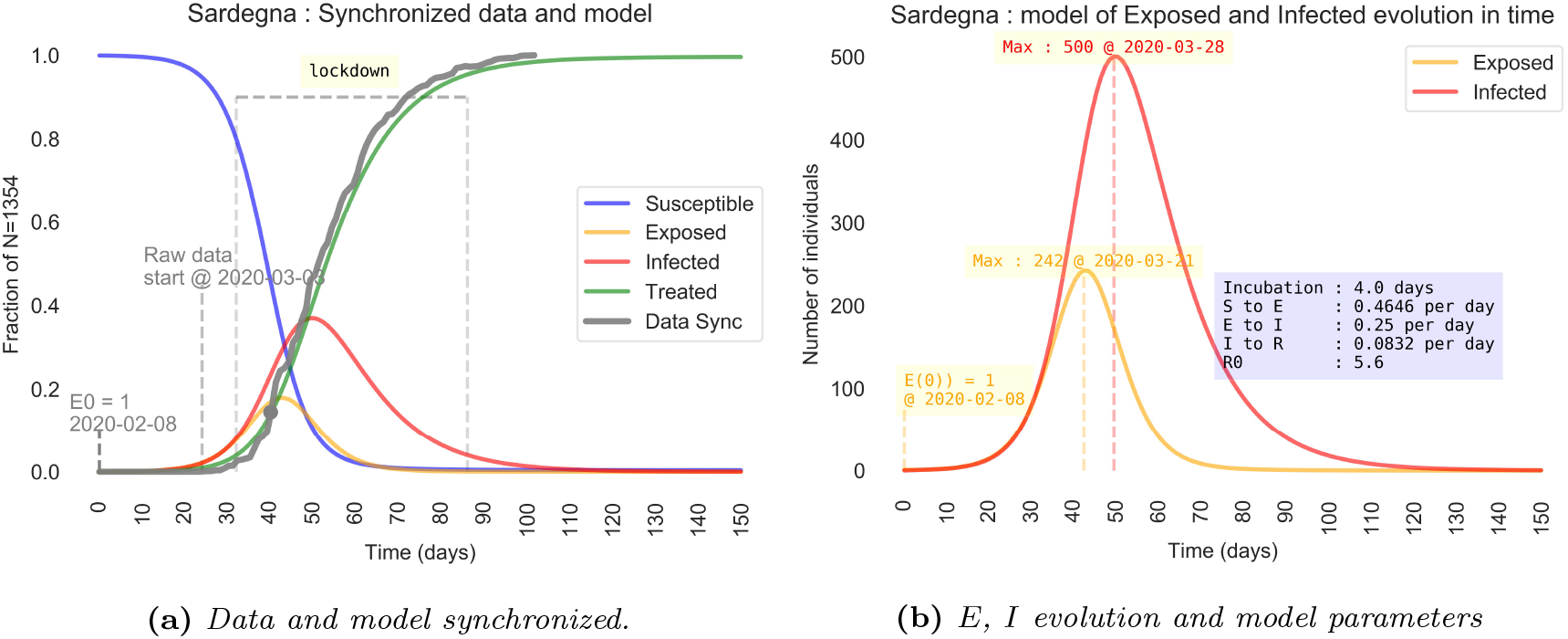
(a) The curve T of the model and T_data_ data has been synchronized at the point where the growth rate has been estimated. The ordinate scale is the ratio on the total number of cases N. (b) Detail of time evolution of E and I curves. The ordinate scale is the the number of individuals in the compartments E, I.

**Figure 17:**
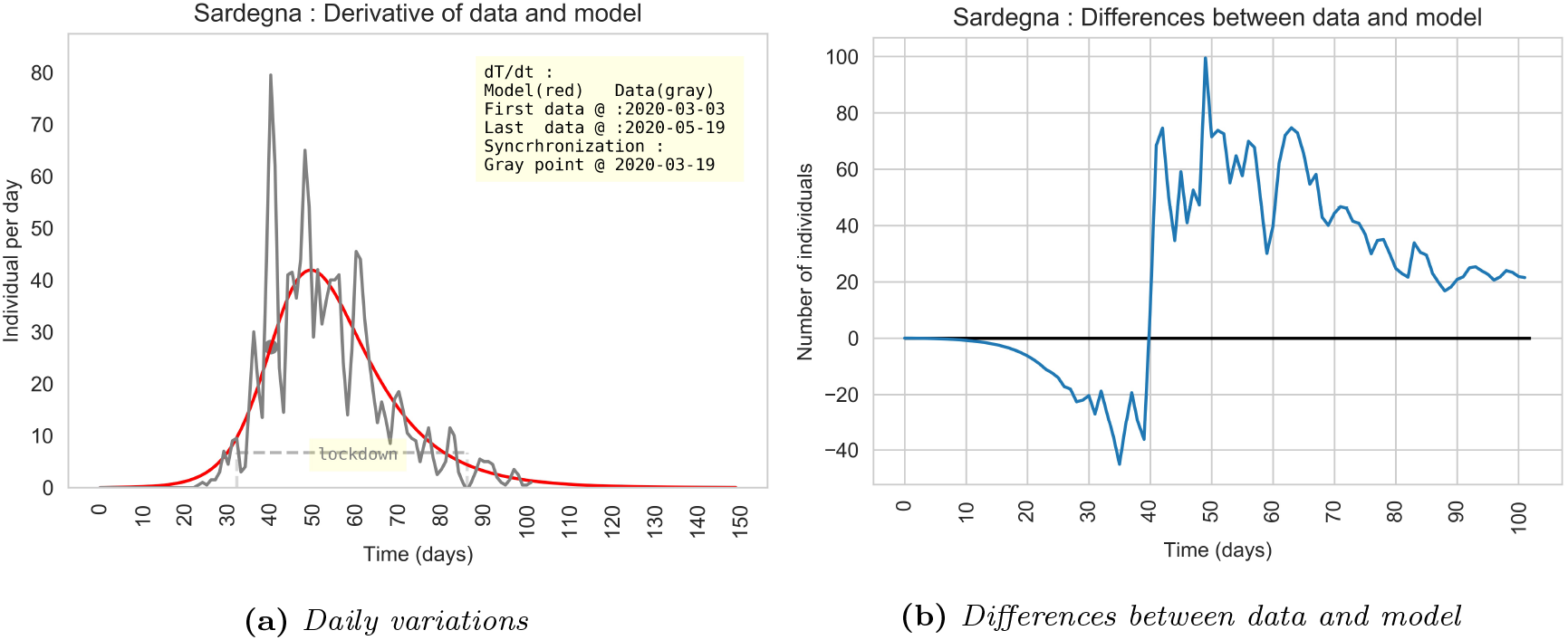
(a) Daily variation of T_data_ and of the T model (red). (b) Differences between T_data_ and the model T.

## Data Availability

Public data from Dipartimento della Protezione Civile Italia.

https://github.com/pcm-dpc/COVID-19/blob/master/dati-regioni/dpc-covid19-ita-regioni.csv

http://opendatadpc.maps.arcgis.com/apps/opsdashboard/index.html#/b0c68bce2cce478eaac82fe38d4138b1

